# Protection of Omicron sub-lineage infection against reinfection with another Omicron sub-lineage

**DOI:** 10.1101/2022.02.24.22271440

**Authors:** Hiam Chemaitelly, Houssein H. Ayoub, Peter Coyle, Patrick Tang, Hadi M. Yassine, Hebah A. Al-Khatib, Maria K. Smatti, Mohammad R. Hasan, Zaina Al-Kanaani, Einas Al-Kuwari, Andrew Jeremijenko, Anvar Hassan Kaleeckal, Ali Nizar Latif, Riyazuddin Mohammad Shaik, Hanan F. Abdul-Rahim, Gheyath K. Nasrallah, Mohamed Ghaith Al-Kuwari, Adeel A. Butt, Hamad Eid Al-Romaihi, Mohamed H. Al-Thani, Abdullatif Al-Khal, Roberto Bertollini, Laith J. Abu-Raddad

## Abstract

**BACKGROUND:** The SARS-CoV-2 Omicron (B.1.1.529) variant has two main sub-lineages, BA.1 and BA.2 with significant genetic distance between them. This study investigated protection of infection with one sub-lineage against reinfection with the other sub-lineage in Qatar during a large BA.1 and BA.2 Omicron wave, from December 19, 2021 to February 21, 2022.

**METHODS:** Two national matched, retrospective cohort studies were conducted to estimate effectiveness of BA.1 infection against reinfection with BA.2 (N=20,197; BA.1-against-BA.2 study), and effectiveness of BA.2 infection against reinfection with BA.1 (N=100,925; BA.2-against-BA.1 study). Associations were estimated using Cox proportional-hazards regression models.

**RESULTS:** In the BA.1-against-BA.2 study, cumulative incidence of infection was estimated at 0.03% (95% CI: 0.01-0.07%) for the BA.1-infected cohort and at 0.62% (95% CI: 0.51-0.75%) for the uninfected-control cohort, 15 days after the start of follow-up. Effectiveness of BA.1 infection against reinfection with BA.2 was estimated at 94.9% (95% CI: 88.4-97.8%). In the BA.2-against-BA.1 study, cumulative incidence of infection was estimated at 0.03% (95% CI: 0.02-0.04%) for the BA.2-infected cohort and at 0.17% (95% CI: 0.15-0.21%) for the uninfected-control cohort, 15 days after the start of follow-up. Effectiveness of BA.2 infection against reinfection with BA.1 was estimated at 85.6% (95% CI: 77.4-90.9%).

**CONCLUSIONS:** Infection with an Omicron sub-lineage appears to induce strong, but not full protection against reinfection with the other sub-lineage, for at least several weeks after the initial infection.

## Introduction

Reinfections with the severe acute respiratory syndrome coronavirus 2 (SARS-CoV-2) variants that can evade immune response are a concern, potentially challenging the global response to the pandemic.^1^ This is especially true of the Omicron^2^ (B.1.1.529) variant and its two main sub-lineages, BA.1 and BA.2, which harbor multiple mutations that can mediate immune evasion.^2-4^ While SARS-CoV-2 infection with earlier variants elicits >85% protection against reinfection with the Alpha^2^ (B.1.1.7),^5-8^ Beta^2^ (B.1.351),^5,7,8^ and Delta^2^ (B.1.617.2)^7,9^ variants, protection against reinfection with the Omicron BA.1 sub-lineage is inferior at <60%.^7^

Qatar has been experiencing a large Omicron wave that started on December 19, 2021 and peaked in mid-January, 2022.^7,10-12^ Initially, the BA.1 sub-lineage was predominant, but within days, the BA.2 sub-lineage predominated (Figure 1). Considering the significant genetic distance between BA.1 and BA.2, we aimed to investigate and estimate protection of prior infection with each sub-lineage against the other.

**Figure 1.**
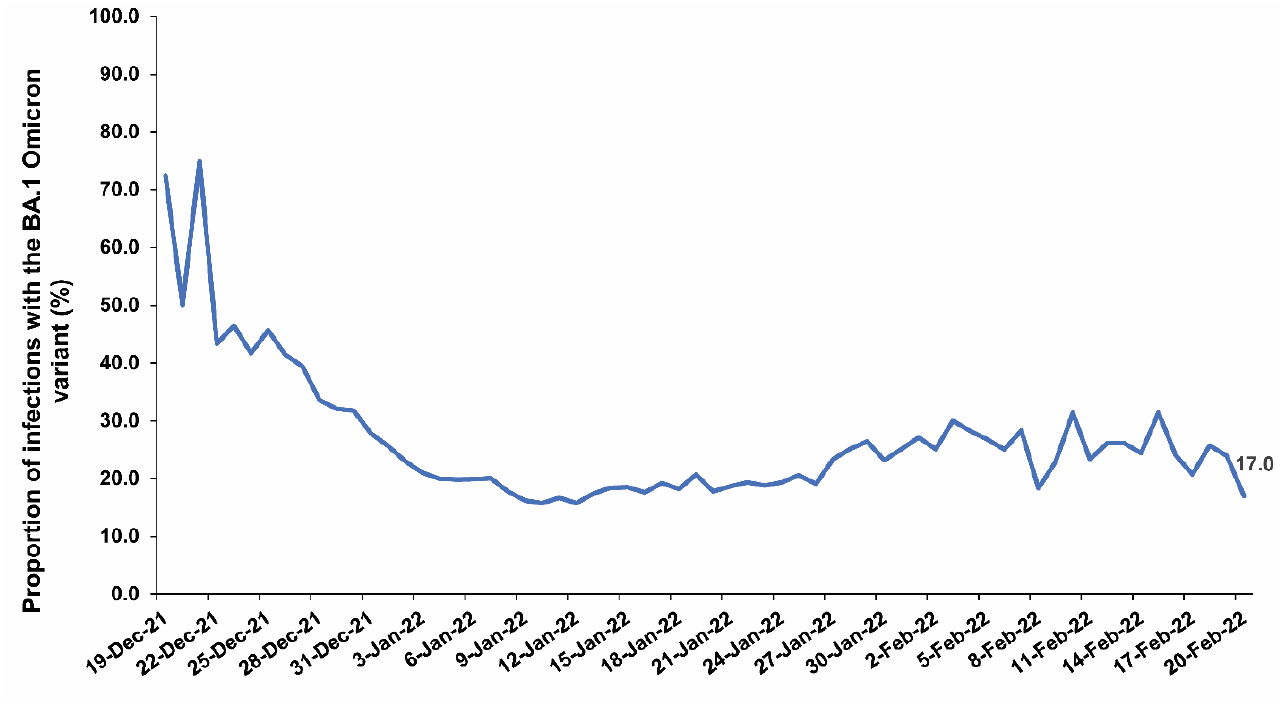
Proportion of BA.1 (versus BA.2) Omicron infections in PCR-positive tests assessed using TaqPath COVID-19 Combo Kit during the study period.

## Methods

### Data sources and study design

This study analyzed the national, federated databases for coronavirus disease 2019 (COVID-19), retrieved from the integrated nationwide digital-health information platform. Databases include all SARS-CoV-2-related data and associated demographic information, with no missing information, since pandemic onset. These include all polymerase chain reaction (PCR) testing and more recently, rapid antigen testing (RAT) conducted at healthcare facilities (from January 5, 2022 onwards). These also include all COVID-19 vaccination records, COVID-19 hospitalizations, infection severity and mortality classifications per World Health Organization (WHO) guidelines,^13,14^ in addition to sex, age, and nationality information retrieved from the national registry. Further description of these national databases can be found in previous publications.^5,15-18^

During this study, from December 19, 2021 to February 21, 2022, nearly all infection incidence was due to the Omicron variant. A total of 315 random SARS-CoV-2-positive specimens collected between December 19, 2021 and January 22, 2022 were viral whole-genome sequenced. Of these, 300 (95.2%) were confirmed as Omicron infections and 15 (4.8%) as Delta infections.^7,10-12^ Of 286 Omicron infections with confirmed sub-lineage status, 68 (23.8%) were BA.1 cases and 218 (76.2%) were BA.2 cases. No Delta case was detected in sequencing after January 8, 2022, nor were other variants. Further details about viral genome sequencing and real-time, reverse-transcription PCR (RT-qPCR) genotyping of random specimens during the study are found in Supplementary Appendix Section S1.

Informed by viral genome sequencing and RT-qPCR genotyping, a SARS-CoV-2 infection with the BA.1 sub-lineage was proxied as an S-gene “target failure” (SGTF) case using the TaqPath COVID-19 Combo Kit (Thermo Fisher Scientific, USA).^19^ Conversely, an infection with the BA.2 sub-lineage was proxied as a non-SGTF case using the same assay.

We assessed effectiveness of BA.1 infection against reinfection with BA.2 (denoted as 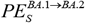 ; BA.1-against-BA.2 study), and effectiveness of BA.2 infection against reinfection with BA.1 (denoted as 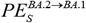; BA.2-against-BA.1 study), using two matched, retrospective cohort studies. *PE*_*S*_ was defined as the proportional reduction in susceptibility to documented infection, regardless of symptoms, among those with the prior sub-lineage infection versus those without.^7,8^ The BA.1-against-BA.2 study followed a cohort of individuals with documented BA.1 infections and compared incidence of BA.2 infection in this cohort with that in a control cohort of individuals with no record of prior SARS-CoV-2 infection. The BA.2-against-BA.1 study followed a cohort of individuals with documented BA.2 infections and compared incidence of BA.1 infection in this cohort with that in a control cohort of individuals with no record of prior SARS-CoV-2 infection.

To optimize specificity in defining the cohorts, the BA.1-infected and BA.2-infected cohorts were defined based on existence of an infection documented only using PCR and with a PCR cycle threshold value <30, between December 19, 2021 and February 21, 2022. In all cohorts of the two studies, the two case and two control cohorts, persons with a record of a prior infection before December 19, 2021 were excluded. This is to ensure that estimated 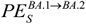 and 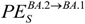 are not affected by immunity induced by prior infections with earlier variants. Record of COVID-19 vaccination was not an exclusion criterion, but the regression analyses adjusted for vaccination status (unvaccinated, one dose, two doses, or three doses at the start of the follow-up). The control cohorts in the two studies were defined on the basis of PCR-negative tests between November 1, 2021 and December 18, 2021 (Figure 2), to ensure that all persons in these cohorts have a record of a recent active residence in Qatar.

**Figure 2.**
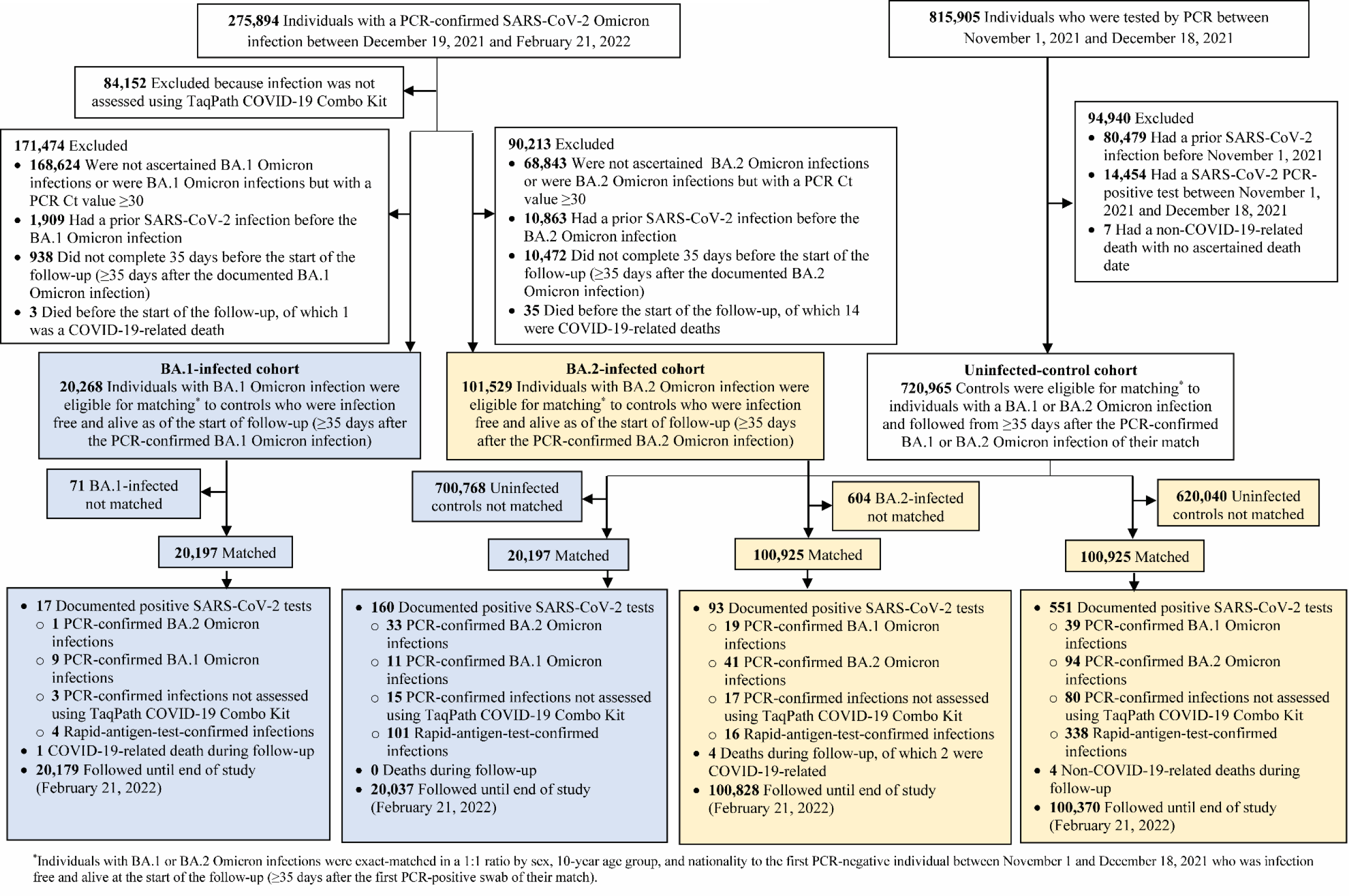
Cohort selection in the BA.1-against-BA.2 and BA.2-against-BA.1 studies.

Ideally, SARS-CoV-2 reinfection is defined as a documented infection ≥90 days after an earlier infection, to avoid misclassification of prolonged infections as reinfections, if a shorter time interval is used.^20-22^ Since the Omicron wave started only few weeks ago, this definition could not be used. Analysis of durations of follow-up was conducted to identify the longest time interval possible while maintaining adequate durations of follow-up and precision of estimates. Informed by the analysis, reinfection was defined as a documentation of infection ≥35 days after the prior infection. At this interval, only a small number of documented reinfections could have been prolonged prior infections rather than true reinfections.^20-24^ Cohorts were thus followed after completion of 35 days since documentation of the BA.1 (or BA.2) infection.

Individuals in each of the BA.1-infected and BA.2-infected cohorts were exact-matched in a 1:1 ratio by sex, 10-year age group, and nationality to uninfected individuals in control cohorts (Figure 2), to control for known differences in the risk of exposure to SARS-CoV-2 infection in Qatar.^15,25-28^ Matching was performed through an iterative process that ensured that each control was alive and infection-free at the start of follow-up. Follow-up was defined, for each matched pair, at ≥35 days after documentation of the BA.1 infection in the BA.1-infected cohort and BA.2 infection in the BA.2-infected cohort. Cohorts were followed up until the first of the following events: a PCR-documented BA.1 infection, a PCR-documented BA.2 infection, other PCR-documented infection (documented with an assay other than TaqPath), RAT-documented infection, death, and end of study censoring (February 21, 2022).

### Laboratory methods

Laboratory methods for the RT-qPCR testing, rapid antigen testing, and viral genome sequencing are found in Section S1.

### Statistical analysis

Frequency distributions and measures of central tendency were used to describe full and matched cohorts. Group comparisons were performed using standardized mean differences (SMDs), with an SMD <0.1 indicating adequate matching.^29^ Cumulative incidence of infection was defined as the proportion of individuals at risk whose primary endpoint was an incident infection during follow-up, and was estimated in each cohort using the Kaplan–Meier estimator method.^30^ Incidence rate of infection in each cohort, which was defined as the number of identified infections divided by the number of person-weeks contributed by all individuals in the cohort, was estimated, along with its 95% confidence interval (CI), using a Poisson log-likelihood regression model with the STATA 17.0 *stptime* command.

The hazard ratio comparing incidence of infection in case versus control cohorts and corresponding 95% CI were calculated using Cox regression adjusted for matching factors and COVID-19 vaccination status (unvaccinated, one dose, two doses, or three doses at the start of the follow-up) with the STATA 17.0 *stcox* command. Shoenfeld residuals and log-log plots for survival curves were used to test the proportional-hazards assumption and to investigate its adequacy. 95% CIs were not adjusted for multiplicity and should not be used to infer definitive differences between cohorts. Interactions were not considered. 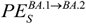 and 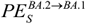were estimated using the equation: Effectiveness = 1−adjusted hazrad ratio. Statistical analyses were conducted in STATA/SE version 17.0.

Due to the large Omicron wave, use of rapid antigen testing was expanded rapidly to supplement PCR testing starting from January 5, 2022, precluding ascertainment of the Omicron sub-lineage in these tests. While 70.2% of all PCR tests (positive or negative) during the study were conducted using an assay that targets the S-gene, a minority of infections were documented with other commercial PCR kits/platforms that are not affected by the del69/70 mutation in the S-gene (Section S1), also precluding ascertainment of the Omicron sub-lineage in these tests.

The main estimate for each of 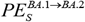 and 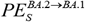 was generated by randomly assigning a sub-lineage status (BA.1 or BA.2) for each RAT-documented infection and non-TaqPath PCR-documented infection, diagnosed in a specific calendar day, on the basis of the probability of the infection being BA.1 or BA.2 in that specific day. This probability was determined by the observed distribution of identified BA.1 and BA.2 infections in each calendar day (Figure 1). 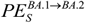 was then estimated by setting the study outcome in the analysis of the BA.1-against-BA.2 study as a BA.2 infection. 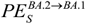 was estimated by setting the study outcome in the analysis of the BA.2-against-BA.1 study as a BA.1 infection.

In a sensitivity analysis, a second estimate was provided for each of 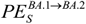 and 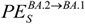, by randomly assigning a sub-lineage status (BA.1 or BA.2) for each RAT-documented infection and non-TaqPath PCR-documented infection, in each cohort of the two studies, on the basis of the observed distribution of identified BA.1 and BA.2 infections among the infections diagnosed in the specific considered cohort.

### Oversight

Hamad Medical Corporation and Weill Cornell Medicine-Qatar Institutional Review Boards approved this retrospective study with a waiver of informed consent. The study was reported following STROBE guidelines. The STROBE checklist is found in Table S1.

## Results

### BA.1-against-BA.2 study

Figure 2 shows the population selection process for the BA.1-against-BA.2 study. Table 1 shows baseline characteristics of full and matched cohorts. The study was based on the total population of Qatar and is broadly representative of the diverse (international), but young and predominantly male, total population of Qatar (Table S2).

**Table 1.** Baseline characteristics of full and matched cohorts in the BA.1-against-BA.2 and BA.2-against-BA.1 studies.

| Characteristics | BA.1-against-BA.2 study |  |  |  |  |  | BA.2-against-BA.2 study |  |  |  |  |  |
| --- | --- | --- | --- | --- | --- | --- | --- | --- | --- | --- | --- | --- |
|  | Full eligible cohorts |  |  | Matched cohorts* |  |  | Full eligible cohorts |  |  | Matched cohorts* |  |  |
|  | BA.1-infected cohort | Uninfected-control cohort | SMD† | BA.1-infected cohort | Uninfected-control cohort | SMD† | BA.2-infected cohort | Uninfected-control cohort | SMD† | BA.2-infected cohort | Uninfected-control cohort | SMD† |
|  | N=20,268 | N=720,965 |  | N=20,197 | N=20,197 |  | N=101,529 | N=720,965 |  | N=100,925 | N=100,925 |  |
| Median age (IQR) — years | 33 (25-41) | 33 (24-41) | 0.07‡ | 33 (25-41) | 33 (24-42) | 0.01‡ | 34 (27-42) | 33 (24-41) | 0.16‡ | 34 (26-42) | 34 (26-43) | 0.00‡ |
| Age group — no. (%) |  |  |  |  |  |  |  |  |  |  |  |  |
| 0-19 years | 3,463 (17.1) | 134,856 (18.7) |  | 3,448 (17.1) | 3,448 (17.1) |  | 13,484 (13.3) | 134,856 (18.7) |  | 13,388 (13.3) | 13,388 (13.3) |  |
| 20-29 years | 4,377 (21.6) | 154,465 (21.4) |  | 4,359 (21.6) | 4,359 (21.6) |  | 21,159 (20.8) | 154,465 (21.4) |  | 21,020 (20.8) | 21,020 (20.8) |  |
| 30-39 years | 6,430 (31.7) | 222,959 (30.9) |  | 6,414 (31.8) | 6,414 (31.8) |  | 34,356 (33.8) | 222,959 (30.9) |  | 34,199 (33.9) | 34,199 (33.9) |  |
| 40-49 years | 3,501 (17.3) | 125,106 (17.4) | 0.05 | 3,495 (17.3) | 3,495 (17.3) | 0.00 | 18,503 (18.2) | 125,106 (17.4) | 0.16 | 18,413 (18.2) | 18,413 (18.2) | 0.00 |
| 50-59 years | 1,676 (8.3) | 57,487 (8.0) |  | 1,668 (8.3) | 1,668 (8.3) |  | 9,457 (9.3) | 57,487 (8.0) |  | 9,403 (9.3) | 9,403 (9.3) |  |
| 60-69 years | 623 (3.1) | 20,571 (2.9) |  | 619 (3.1) | 619 (3.1) |  | 3,347 (3.3) | 20,571 (2.9) |  | 3,307 (3.3) | 3,307 (3.3) |  |
| 70+ years | 198 (1.0) | 5,521 (0.8) |  | 194 (1.0) | 194 (1.0) |  | 1,223 (1.2) | 5,521 (0.8) |  | 1,195 (1.2) | 1,195 (1.2) |  |
| Sex |  |  |  |  |  |  |  |  |  |  |  |  |
| Male | 10,361 (51.1) | 488,631 (67.8) | 0.34 | 10,331 (51.2) | 10,331 (51.2) | 0.00 | 60,886 (60.0) | 488,631 (67.8) | 0.16 | 60,548 (60.0) | 60,548 (60.0) | 0.00 |
| Female | 9,907 (48.9) | 232,334 (32.2) |  | 9,866 (48.9) | 9,866 (48.9) |  | 40,643 (40.0) | 232,334 (32.2) |  | 40,377 (40.0) | 40,377 (40.0) |  |
| Nationality§ |  |  |  |  |  |  |  |  |  |  |  |  |
| Bangladeshi | 301 (1.5) | 49,016 (6.8) |  | 300 (1.5) | 300 (1.5) |  | 3,002 (3.0) | 49,016 (6.8) |  | 2,995 (3.0) | 2,995 (3.0) |  |
| Egyptian | 876 (4.3) | 29,782 (4.1) |  | 873 (4.3) | 873 (4.3) |  | 5,143 (5.1) | 29,782 (4.1) |  | 5,117 (5.1) | 5,117 (5.1) |  |
| Filipino | 2,543 (12.6) | 39,350 (5.5) |  | 2,538 (12.6) | 2,538 (12.6) |  | 13,064 (12.9) | 39,350 (5.5) |  | 13,011 (12.9) | 13,011 (12.9) |  |
| Indian | 2,927 (14.4) | 208,042 (28.9) |  | 2,923 (14.5) | 2,923 (14.5) |  | 20,376 (20.1) | 208,042 (28.9) |  | 20,317 (20.1) | 20,317 (20.1) |  |
| Nepalese | 455 (2.2) | 52,795 (7.3) | 0.72 | 455 (2.3) | 455 (2.3) | 0.00 | 4,565 (4.5) | 52,795 (7.3) | 0.47 | 4,560 (4.5) | 4,560 (4.5) | 0.00 |
| Pakistani | 471 (2.3) | 38,329 (5.3) |  | 470 (2.3) | 470 (2.3) |  | 2,917 (2.9) | 38,329 (5.3) |  | 2,909 (2.9) | 2,909 (2.9) |  |
| Qatari | 6,232 (30.8) | 95,364 (13.2) |  | 6,197 (30.7) | 6,197 (30.7) |  | 23,160 (22.8) | 95,364 (13.2) |  | 22,946 (22.7) | 22,946 (22.7) |  |
| Sri Lankan | 302 (1.5) | 19,812 (2.8) |  | 302 (1.5) | 302 (1.5) |  | 2,788 (2.8) | 19,812 (2.8) |  | 2,782 (2.8) | 2,782 (2.8) |  |
| Sudanese | 612 (3.0) | 12,169 (1.7) |  | 612 (3.0) | 612 (3.0) |  | 3,160 (3.1) | 12,169 (1.7) |  | 3,149 (3.1) | 3,149 (3.1) |  |
| Other nationalities¶ | 5,549 (27.4) | 176,306 (24.5) |  | 5,527 (27.4) | 5,527 (27.4) |  | 23,354 (23.0) | 176,306 (24.5) |  | 23,139 (22.9) | 23,139 (22.9) |  |

Abbreviations: IQR, interquartile range; SMD, standardized mean difference.
\*Cohorts were matched one-to-one by sex, 10-year age group, and nationality.
†SMD is the difference in the mean of a covariate between groups divided by the pooled standard deviation. An SMD <0.1 indicates adequate matching.
‡SMD is for the mean difference between groups divided by the pooled standard deviation.
§Nationalities were chosen to represent the most populous groups in Qatar.
¶These comprise 131 other nationalities in the BA.1-infected cohort and 202 other nationalities in the uninfected-control cohort in the full eligible cohorts of the BA.1-against-BA.2 study, and 129 other nationalities in the BA.1-infected cohort and 129 other nationalities in the uninfected-control cohort in the matched cohorts of the BA.1-against-BA.2 study. These comprise 155 other nationalities in the BA.2-infected cohort and 202 other nationalities in the uninfected-control in the full eligible cohorts of the BA.2-against-BA.1 study, and 151 other nationalities in the BA.2-infected cohort and 151 other nationalities in the uninfected-control in the matched cohorts of the BA.2-against-BA.1 study.

The median time of follow-up was 14 days (interquartile range (IQR), 12-17 days) for the BA.1-infected and 14 days (interquartile range (IQR), 11-17 days) for the uninfected-control cohorts (Figure 3). The proportion of individuals who had a PCR or RAT test during follow-up was 9.5% for the BA.1-infected cohort and 12.2% for the uninfected-control cohort.

**Figure 3.**
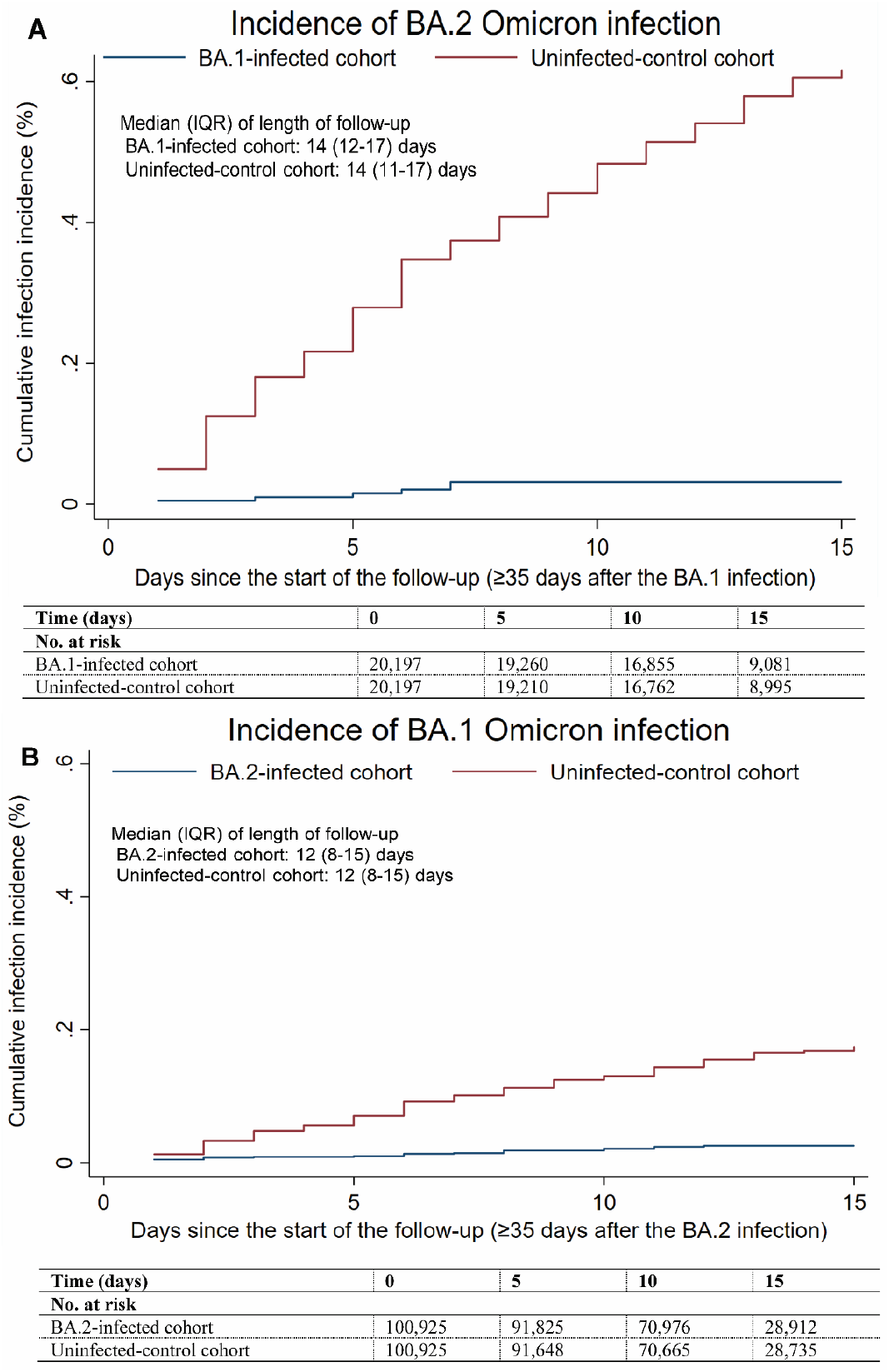
Cumulative incidence of A) BA.2 and B) BA.1 Omicron infections in the BA.1-against-BA.2 and BA.2-against-BA.1 studies, respectively.

One PCR-documented BA.2 infection, 9 PCR-documented BA.1 infections, 3 other PCR-documented infections, and 4 RAT-documented infections were recorded in the BA.1-infected cohort ≥35 days after the BA.1 infection (Figure 2). Thirty-three PCR-documented BA.2 infections, 11 PCR-documented BA.1 infections, 15 other PCR-documented infections, and 101 RAT-documented infections were recorded during the corresponding time of follow-up for the uninfected-control cohort.

In the main analysis for 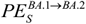, cumulative incidence of infection was estimated at 0.03% (95% CI: 0.01-0.07%) for the BA.1-infected cohort and at 0.62% (95% CI: 0.51-0.75%) for the uninfected-control cohort, 15 days after the start of follow-up (Figure 3). The hazard ratio for infection, adjusted for sex, 10-year age group, nationality group, and vaccination status, was estimated at 0.05 (95% CI: 0.02-0.12) (Table 2). The effectiveness of BA.1 infection against reinfection with BA.2 was estimated at 94.9% (95% CI: 88.4-97.8%). In the sensitivity analysis (Table 2 and Figure S1), the effectiveness of BA.1 infection against reinfection with BA.2 was estimated at 98.4% (95% CI: 93.3-99.6%).

**Table 2.**
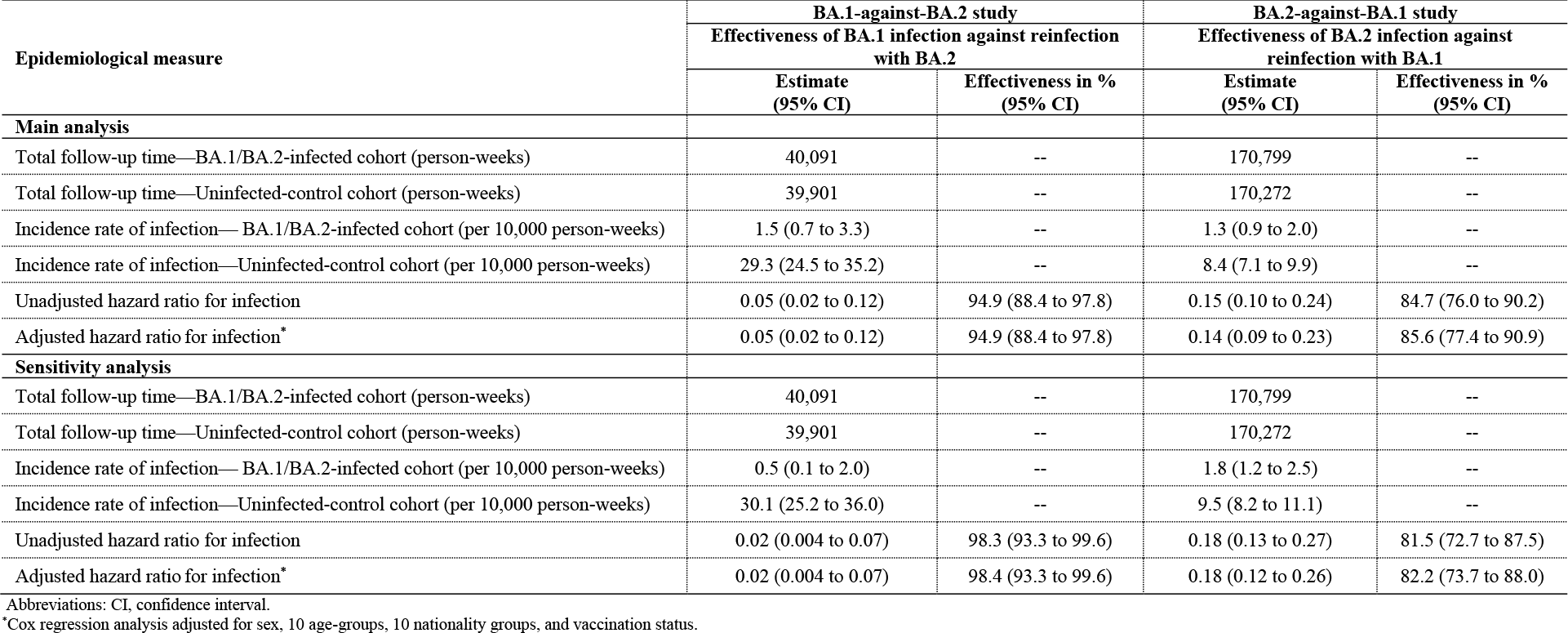
Effectiveness against reinfection in the BA.1-against-BA.2 and BA.2-against-BA.1 studies.

| Epidemiological measure | BA.1-against-BA.2 study |  | BA.2-against-BA.1 study |  |
| --- | --- | --- | --- | --- |
|  | Effectiveness of BA.1 infection against reinfection with BA.2 |  | Effectiveness of BA.2 infection against reinfection with BA.1 |  |
|  | Estimate (95% CI) | Effectiveness in % (95% CI) | Estimate (95% CI) | Effectiveness in % (95% CI) |
| **Main analysis** |  |  |  |  |
| Total follow-up time—BA.1/BA.2-infected cohort (person-weeks) | 40,091 | -- | 170,799 | -- |
| Total follow-up time—Uninfected-control cohort (person-weeks) | 39,901 | -- | 170,272 | -- |
| Incidence rate of infection— BA.1/BA.2-infected cohort (per 10,000 person-weeks) | 1.5 (0.7 to 3.3) | -- | 1.3 (0.9 to 2.0) | -- |
| Incidence rate of infection—Uninfected-control cohort (per 10,000 person-weeks) | 29.3 (24.5 to 35.2) | -- | 8.4 (7.1 to 9.9) | -- |
| Unadjusted hazard ratio for infection | 0.05 (0.02 to 0.12) | 94.9 (88.4 to 97.8) | 0.15 (0.10 to 0.24) | 84.7 (76.0 to 90.2) |
| Adjusted hazard ratio for infection* | 0.05 (0.02 to 0.12) | 94.9 (88.4 to 97.8) | 0.14 (0.09 to 0.23) | 85.6 (77.4 to 90.9) |
| **Sensitivity analysis** |  |  |  |  |
| Total follow-up time—BA.1/BA.2-infected cohort (person-weeks) | 40,091 | -- | 170,799 | -- |
| Total follow-up time—Uninfected-control cohort (person-weeks) | 39,901 | -- | 170,272 | -- |
| Incidence rate of infection— BA.1/BA.2-infected cohort (per 10,000 person-weeks) | 0.5 (0.1 to 2.0) | -- | 1.8 (1.2 to 2.5) | -- |
| Incidence rate of infection—Uninfected-control cohort (per 10,000 person-weeks) | 30.1 (25.2 to 36.0) | -- | 9.5 (8.2 to 11.1) | -- |
| Unadjusted hazard ratio for infection | 0.02 (0.004 to 0.07) | 98.3 (93.3 to 99.6) | 0.18 (0.13 to 0.27) | 81.5 (72.7 to 87.5) |
| Adjusted hazard ratio for infection* | 0.02 (0.004 to 0.07) | 98.4 (93.3 to 99.6) | 0.18 (0.12 to 0.26) | 82.2 (73.7 to 88.0) |

Abbreviations: CI, confidence interval.
\*Cox regression analysis adjusted for sex, 10 age-groups, 10 nationality groups, and vaccination status.

### BA.2-against-BA.1 study

Figure 2 shows the population selection process for the BA.2-against-BA.1 study. Table 1 shows baseline characteristics of full and matched cohorts. The study population was representative of the population of Qatar (Table S2).

The median time of follow-up was 12 days (IQR, 8-15 days) for both the BA.2-infected cohort and the uninfected-control cohort (Figure 3). The proportion of individuals who had a PCR or RAT test during follow-up was 6.4% for the BA.2-infected cohort and 8.8% for the uninfected-control cohort.

Nineteen PCR-documented BA.1 infections, 41 PCR-documented BA.2 infections, 17 other PCR-documented infections, and 16 RAT-documented infections were recorded in the BA.2-infected cohort ≥35 days after the BA.2 infection (Figure 2). Thirty-nine PCR-documented BA.1 infections, 94 PCR-documented BA.2 infections, 80 other PCR-documented infections, and 338 RAT-documented infections were recorded during the corresponding time of follow-up for the uninfected-control cohort.

In the main analysis for 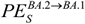, cumulative incidence of infection was estimated at 0.03% (95% CI: 0.02-0.04%) for the BA.2-infected cohort and at 0.17% (95% CI: 0.15-0.21%) for the uninfected-control cohort, 15 days after the start of follow-up (Figure 3). The adjusted hazard ratio for infection was estimated at 0.14 (95% CI: 0.09-0.23) (Table 2). The effectiveness of BA.2 infection against reinfection with BA.1 was estimated at 85.6% (95% CI: 77.4-90.9%). In the sensitivity analysis (Table 2 and Figure S1), the effectiveness of BA.2 infection against reinfection with BA.1 was estimated at 82.2% (95% CI: 73.7-88.0%).

## Discussion

Reinfections with BA.2 (or BA.1) shortly after infection with BA.1 (or BA.2) have been observed in Qatar during a large Omicron wave in which both sub-lineages were intensely circulating. Indeed, 0.9% (1,062 cases) of all individuals who had a PCR-positive test with a known sub-lineage status during the Omicron wave, between December 19, 2021 and February 21, 2022, had also a subsequent PCR-positive test with the other counter sub-lineage within the same duration.

However, it is remarkable that incidence of reinfection, regardless of sub-lineage, was much lower in the BA.1-infected and BA.2-infected cohorts than incidence of infection in the corresponding uninfected-control cohorts (Figure 2), consistent with strong protection against reinfection regardless of sub-lineage. Our findings indicate that infection with an Omicron sub-lineage appears to elicit strong protection against reinfection with the other sub-lineage at an effectiveness that exceeds 85%, similar to the protection observed for infection with original virus or earlier variants (Alpha, Beta, or Delta) against reinfection with original virus or earlier variants.^2,5-9,20,24^

These findings, in the context of broader evidence for natural immunity,^2,5-9,20-22^ suggest that natural immunity of SARS-CoV-2 variants cluster into two groups: early non-Omicron variants, and Omicron BA.1 and BA.2 sub-lineages. Within each group, there appears to be strong protection against reinfection with an effectiveness that exceeds 85%. However, across groups, the protection may not exceed 60%, as was observed recently.^7^ This conclusion is also supported by evidence on sensitivity of variants to SARS-CoV-2 antibodies.^2-4,31,32^

This study has limitations. Since the Omicron wave was initially dominated with BA.1 (Figure 1), the follow-up in the BA.2-against-BA.1 study was shifted in calendar time to after the follow-up in the BA.1-against-BA.2 study. With the high intensity of infection transmission, followed by rapid decline of the Omicron wave, more of the uninfected-controls in the BA.2-against-BA.1 study may have experienced an *undocumented* Omicron infection compared to the uninfected-controls in the BA.1-against-BA.2 study. This would bias 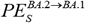 to a lower value and may explain why 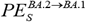 was lower than that of 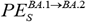.

Effectiveness against reinfection was estimated for only few weeks after the primary infection. A longer duration of follow-up may identify differences not yet seen given the recency of the Omicron wave. However, evidence has been consistent that natural immunity, unlike vaccine immunity, wanes slowly with minimal waning for at least several months after primary infection.^2,5-9,20-22^

BA.1 and BA.2 ascertainment was based on proxy criteria, presence or absence of SGTF using the TaqPath PCR assay, but this method of ascertainment is well established not only for Omicron sub-lineages, but also for other variants such as Alpha.^6,19,33^ BA.1 and BA.2 ascertainment was not possible for infections diagnosed using RAT or other PCR testing. This limitation was mitigated by basing the main analysis estimate on the distribution of known BA.1 and BA.2 cases for each calendar day, and by providing a sensitivity analysis where the estimate was made based on the distribution of known BA.1 and BA.2 cases in each cohort of the two studies.

Some Omicron infections may have been misclassified Delta infections, but this is not likely, as Delta incidence was limited during the time of follow-up (Section S1). With the recency of the Omicron wave, we had to use a short interval of 35 days to define reinfection, perhaps introducing bias due to misclassification of prolonged infections as reinfections. However, such potential bias is less likely to affect 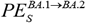and 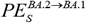, but may affect estimates of effectiveness of BA.1 (or BA.2) infection against reinfection with BA.1 (or BA.2); that is, when both the primary infection and the reinfection are both due to the same sub-lineage. Such effectiveness estimates are not reported in this study (but found in a separate analysis to be comparable to 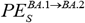 and 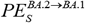). Regardless, such bias leads to underestimation of effectiveness, as it would inflate incident cases only in the BA.1-infected or BA.2 infected cohorts, thereby further supporting our finding of strong protection against reinfection.

As an observational study, the investigated cohorts were neither blinded nor randomized, so unmeasured or uncontrolled confounding cannot be excluded. While matching was done for sex, age, and nationality, this was not possible for other factors, such as comorbidities, occupation, or geography, as such data were not available. However, matching was done to control for factors that affect infection exposure in Qatar.^15,25-28^ Matching by age may have reduced potential bias due to comorbidities. The number of individuals with severe chronic conditions is also small in Qatar’s young population.^15,34^ Matching by nationality may have partially controlled for differences in occupational risk or socio-economic status, given the association between nationality and occupation in Qatar.^15,25-28^ Qatar is essentially a city state and infection incidence was broadly distributed across neighborhoods/areas; that is, geography is not likely to have been a confounding factor. Lastly, matching by the considered factors has been shown to provide adequate control of bias in studies that used control groups in Qatar.^16,35-38^ These control groups included unvaccinated cohorts versus vaccinated cohorts within two weeks of the first dose,^16,35-37^ when vaccine protection is negligible,^39,40^ and mRNA-1273-versus BNT162b2-vaccinated cohorts, also in the first two weeks after the first dose.^38^ A strength of this study is exclusion of those with a documented prior infection before the Omicron wave, to minimize potential confounding introduced by natural immunity due to earlier variants.

In conclusion, infection with an Omicron sub-lineage appears to induce strong, but not full protection against reinfection with the other sub-lineage, for at least several weeks after the initial infection.

## Data Availability

The dataset of this study is a property of the Qatar Ministry of Public Health that was provided to the researchers through a restricted-access agreement that prevents sharing the dataset with a third party or publicly. Future access to this dataset can be considered through a direct application for data access to Her Excellency the Minister of Public Health (https://www.moph.gov.qa/english/Pages/default.aspx). Aggregate data are available within the manuscript and its Supplementary information.

## Sources of support and acknowledgements

We acknowledge the many dedicated individuals at Hamad Medical Corporation, the Ministry of Public Health, the Primary Health Care Corporation, Qatar Biobank, Sidra Medicine, and Weill Cornell Medicine-Qatar for their diligent efforts and contributions to make this study possible. The authors are grateful for institutional salary support from the Biomedical Research Program and the Biostatistics, Epidemiology, and Biomathematics Research Core, both at Weill Cornell Medicine-Qatar, as well as for institutional salary support provided by the Ministry of Public Health, Hamad Medical Corporation, and Sidra Medicine. The authors are also grateful for the Qatar Genome Programme and Qatar University Biomedical Research Center for institutional support for the reagents needed for the viral genome sequencing. The funders of the study had no role in study design, data collection, data analysis, data interpretation, or writing of the article. Statements made herein are solely the responsibility of the authors.

## Author contributions

HC co-designed the study, performed the statistical analyses, and co-wrote the first draft of the article. LJA conceived and co-designed the study, led the statistical analyses, and co-wrote the first draft of the article. HY, HAK, and MS conducted viral genome sequencing. PT and MRH conducted the multiplex, RT-qPCR variant screening and viral genome sequencing. All authors contributed to data collection and acquisition, database development, discussion and interpretation of the results, and to the writing of the manuscript. All authors have read and approved the final manuscript.

## Supplementary Appendix

### Section 1. Laboratory methods and variant ascertainment

#### Real-time reverse-transcription polymerase chain reaction testing

Nasopharyngeal and/or oropharyngeal swabs were collected for PCR testing and placed in Universal Transport Medium (UTM). Aliquots of UTM were: 1) extracted on KingFisher Flex (Thermo Fisher Scientific, USA), MGISP-960 (MGI, China), or ExiPrep 96 Lite (Bioneer, South Korea) followed by testing with real-time reverse-transcription PCR (RT-qPCR) using TaqPath COVID-19 Combo Kits (Thermo Fisher Scientific, USA) on an ABI 7500 FAST (Thermo Fisher Scientific, USA); 2) tested directly on the Cepheid GeneXpert system using the Xpert Xpress SARS-CoV-2 (Cepheid, USA); or 3) loaded directly into a Roche cobas 6800 system and assayed with the cobas SARS-CoV-2 Test (Roche, Switzerland). The first assay targets the viral S, N, and ORF1ab gene regions. The second targets the viral N and E-gene regions, and the third targets the ORF1ab and E-gene regions.

All PCR testing was conducted at the Hamad Medical Corporation Central Laboratory or Sidra Medicine Laboratory, following standardized protocols.

#### Rapid antigen testing

SARS-CoV-2 antigen tests were performed on nasopharyngeal swabs using one of the following lateral flow antigen tests: Panbio COVID-19 Ag Rapid Test Device (Abbott, USA); SARS-CoV-2 Rapid Antigen Test (Roche, Switzerland); Standard Q COVID-19 Antigen Test (SD Biosensor, Korea); or CareStart COVID-19 Antigen Test (Access Bio, USA). All antigen tests were performed point-of-care according to each manufacturer’s instructions at public or private hospitals and clinics throughout Qatar with prior authorization and training by the Ministry of Public Health (MOPH). Antigen test results were electronically reported to the MOPH in real time using the Antigen Test Management System which is integrated with the national COVID-19 database.

#### Classification of infections by variant type

Surveillance for SARS-CoV-2 variants in Qatar is mainly based on viral genome sequencing and multiplex RT-qPCR variant screening^1^ of random positive clinical samples,^2-7^ complemented by deep sequencing of wastewater samples.^4,8^

A total of 315 random SARS-CoV-2-positive specimens collected between December 19, 2021 and January 22, 2022 were viral whole-genome sequenced on a Nanopore GridION sequencing device. Of these, 300 (95.2%) were confirmed as Omicron infections and 15 (4.8%) as Delta (B.1.617.2)^9^ infections.^4,10^ Of 286 Omicron infections with confirmed sub-lineage status, 68 (23.8%) were BA.1 cases and 218 (76.2%) were BA.2 cases. No Delta case was detected in sequencing after January 8, 2022, nor were other variants.

Additionally, a total of 1,315 random SARS-CoV-2-positive specimens collected between December 22, 2021 and January 1, 2022 were RT-qPCR genotyped. The RT-qPCR genotyping identified 1 B.1.617.2-like Delta case, 366 BA.1-like Omicron cases, 898 BA.2-like Omicron cases, and 50 were undetermined cases where the genotype could not be assigned.

The accuracy of the RT-qPCR genotyping was verified against either Sanger sequencing of the receptor-binding domain (RBD) of SARS-CoV-2 surface glycoprotein (S) gene, or by viral whole-genome sequencing on a Nanopore GridION sequencing device. From 147 random SARS-CoV-2-positive specimens all collected in December of 2021, RT-qPCR genotyping was able to assign a genotype in 129 samples. The agreement between RT-qPCR genotyping and sequencing was 100% for Delta (n=82), 100% for Omicron BA.1 (n=18), and 93% for Omicron BA.2 (27 of 29 were correctly assigned to BA.2 and remaining 2 specimens genotyped as BA.2 were B.1.617.2 by sequencing). Of the remaining 18 specimens: 10 failed PCR amplification and sequencing, 8 could not be assigned a genotype by RT-qPCR (4 of 8 were B.1.617.2 by sequencing, and the remaining 4 failed sequencing). All the variant RT-qPCR genotyping was conducted at the Sidra Medicine Laboratory following standardized protocols.

The large Omicron-wave exponential-growth phase in Qatar started on December 19, 2021 and peaked in mid-January, 2022.^4,10,11^ The study duration coincided with the intense Omicron wave where Delta incidence was limited. Accordingly, any PCR or rapid antigen positive test during the study duration, between December 19, 2021 and February 21, 2022, was assumed to be an Omicron infection.

Informed by the viral genome sequencing and the RT-qPCR genotyping, a SARS-CoV-2 infection with the BA.1 sub-lineage was proxied as an S-gene “target failure” (SGTF) case using the TaqPath COVID-19 Combo Kits (Thermo Fisher Scientific, USA^12^).^13^ A SARS-CoV-2 infection with the BA.2 sub-lineage was proxied as a non-SGTF case using the TaqPath assay.

**Table S1.**
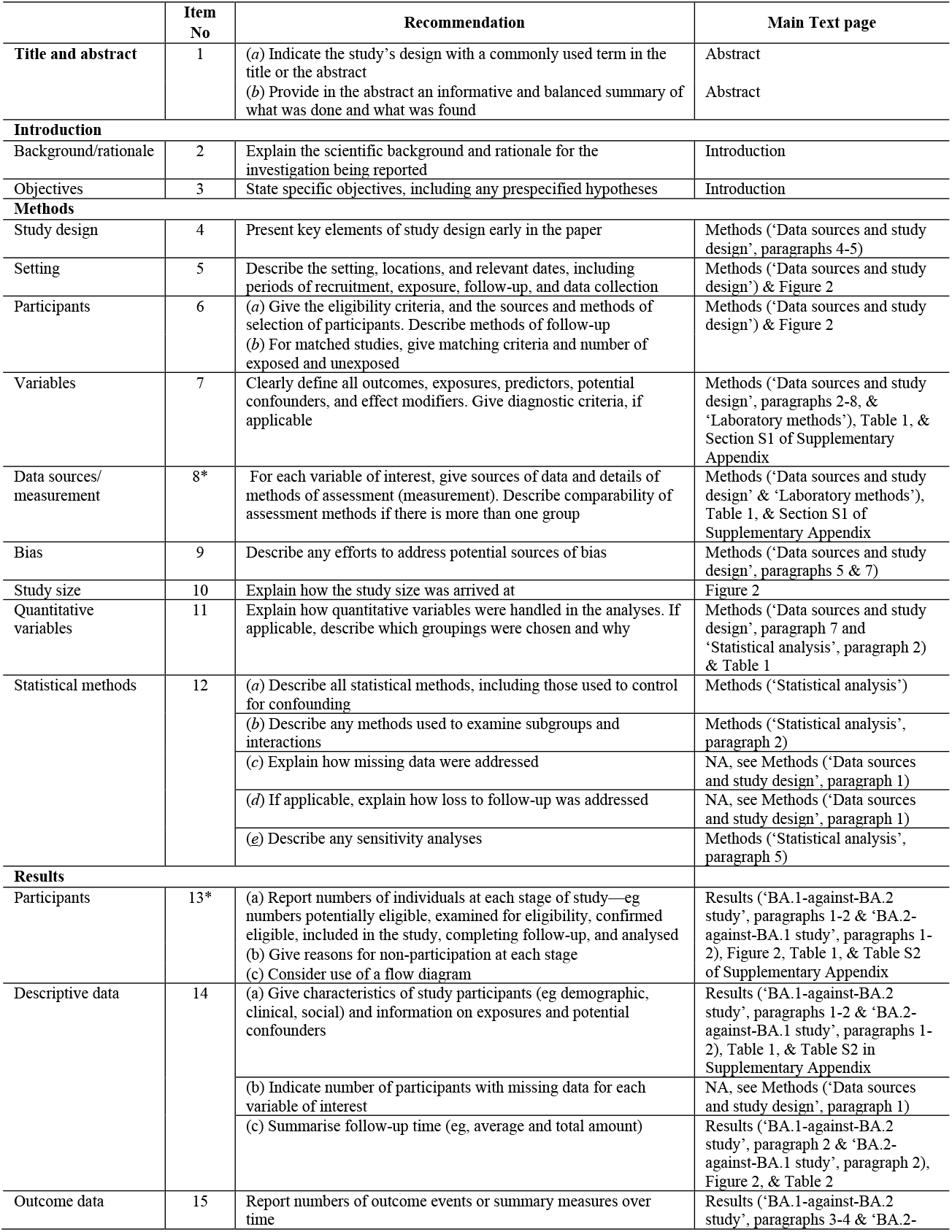
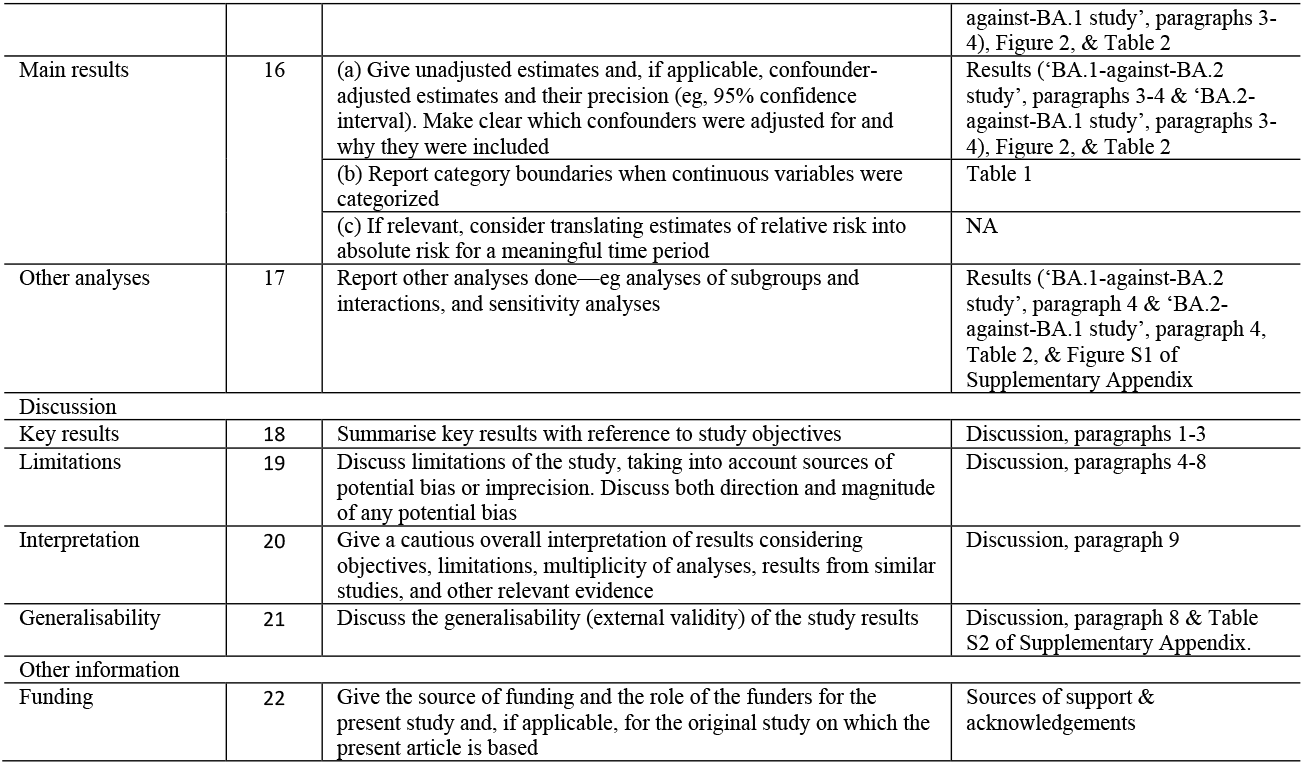
STROBE checklist for cohort studies.

|  | Item No | Recommendation | Main Text page |
| --- | --- | --- | --- |
| **Title and abstract** | 1 | (a) Indicate the study’s design with a commonly used term in the title or the abstract<br>(b) Provide in the abstract an informative and balanced summary of what was done and what was found | Abstract<br>Abstract |
| **Introduction** |  |  |  |
| Background/rationale | 2 | Explain the scientific background and rationale for the investigation being reported | Introduction |
| Objectives | 3 | State specific objectives, including any prespecified hypotheses | Introduction |
| **Methods** |  |  |  |
| Study design | 4 | Present key elements of study design early in the paper | Methods (‘Data sources and study design’, paragraphs 4-5) |
| Setting | 5 | Describe the setting, locations, and relevant dates, including periods of recruitment, exposure, follow-up, and data collection | Methods (‘Data sources and study design’) & Figure 2 |
| Participants | 6 | (a) Give the eligibility criteria, and the sources and methods of selection of participants. Describe methods of follow-up<br>(b) For matched studies, give matching criteria and number of exposed and unexposed | Methods (‘Data sources and study design’) & Figure 2 |
| Variables | 7 | Clearly define all outcomes, exposures, predictors, potential confounders, and effect modifiers. Give diagnostic criteria, if applicable | Methods (‘Data sources and study design’, paragraphs 2-8, & ‘Laboratory methods’), Table 1, & Section S1 of Supplementary Appendix |
| Data sources/measurement | 8* | For each variable of interest, give sources of data and details of methods of assessment (measurement). Describe comparability of assessment methods if there is more than one group | Methods (‘Data sources and study design’ & ‘Laboratory methods’), Table 1, & Section S1 of Supplementary Appendix |
| Bias | 9 | Describe any efforts to address potential sources of bias | Methods (‘Data sources and study design’, paragraphs 5 & 7) |
| Study size | 10 | Explain how the study size was arrived at | Figure 2 |
| Quantitative variables | 11 | Explain how quantitative variables were handled in the analyses. If applicable, describe which groupings were chosen and why | Methods (‘Data sources and study design’, paragraph 7 and ‘Statistical analysis’, paragraph 2) & Table 1 |
| Statistical methods | 12 | (a) Describe all statistical methods, including those used to control for confounding | Methods (‘Statistical analysis’) |
|  |  | (b) Describe any methods used to examine subgroups and interactions | Methods (‘Statistical analysis’, paragraph 2) |
|  |  | (c) Explain how missing data were addressed | NA, see Methods (‘Data sources and study design’, paragraph 1) |
|  |  | (d) If applicable, explain how loss to follow-up was addressed | NA, see Methods (‘Data sources and study design’, paragraph 1) |
|  |  | (e) Describe any sensitivity analyses | Methods (‘Statistical analysis’, paragraph 5) |
| **Results** |  |  |  |
| Participants | 13* | (a) Report numbers of individuals at each stage of study—eg numbers potentially eligible, examined for eligibility, confirmed eligible, included in the study, completing follow-up, and analysed<br>(b) Give reasons for non-participation at each stage<br>(c) Consider use of a flow diagram | Results (‘BA.1-against-BA.2 study’, paragraphs 1-2 & ‘BA.2-against-BA.1 study’, paragraphs 1-2), Figure 2, Table 1, & Table S2 of Supplementary Appendix |
| Descriptive data | 14 | (a) Give characteristics of study participants (eg demographic, clinical, social) and information on exposures and potential confounders | Results (‘BA.1-against-BA.2 study’, paragraphs 1-2 & ‘BA.2-against-BA.1 study’, paragraphs 1-2), Table 1, & Table S2 in Supplementary Appendix |
|  |  | (b) Indicate number of participants with missing data for each variable of interest | NA, see Methods (‘Data sources and study design’, paragraph 1) |
|  |  | (c) Summarise follow-up time (eg, average and total amount) | Results (‘BA.1-against-BA.2 study’, paragraph 2 & ‘BA.2-against-BA.1 study’, paragraph 2), Figure 2, & Table 2 |
| Outcome data | 15 | Report numbers of outcome events or summary measures over time | Results (‘BA.1-against-BA.2 study’, paragraphs 3-4 & ‘BA.2- |
|  |  |  | against-BA.1 study', paragraphs 3-4), Figure 2, & Table 2 |
| Main results | 16 | (a) Give unadjusted estimates and, if applicable, confounder-adjusted estimates and their precision (eg, 95% confidence interval). Make clear which confounders were adjusted for and why they were included | Results ('BA.1-against-BA.2 study', paragraphs 3-4 & 'BA.2-against-BA.1 study', paragraphs 3-4), Figure 2, & Table 2 |
|  |  | (b) Report category boundaries when continuous variables were categorized | Table 1 |
|  |  | (c) If relevant, consider translating estimates of relative risk into absolute risk for a meaningful time period | NA |
| Other analyses | 17 | Report other analyses done—eg analyses of subgroups and interactions, and sensitivity analyses | Results ('BA.1-against-BA.2 study', paragraph 4 & 'BA.2-against-BA.1 study', paragraph 4, Table 2, & Figure S1 of Supplementary Appendix |
| Discussion |  |  |  |
| Key results | 18 | Summarise key results with reference to study objectives | Discussion, paragraphs 1-3 |
| Limitations | 19 | Discuss limitations of the study, taking into account sources of potential bias or imprecision. Discuss both direction and magnitude of any potential bias | Discussion, paragraphs 4-8 |
| Interpretation | 20 | Give a cautious overall interpretation of results considering objectives, limitations, multiplicity of analyses, results from similar studies, and other relevant evidence | Discussion, paragraph 9 |
| Generalisability | 21 | Discuss the generalisability (external validity) of the study results | Discussion, paragraph 8 & Table S2 of Supplementary Appendix. |
| Other information |  |  |  |
| Funding | 22 | Give the source of funding and the role of the funders for the present study and, if applicable, for the original study on which the present article is based | Sources of support & acknowledgements |

**Table S2.** Representativeness of study participants.

| Category |  |
| --- | --- |
| Disease, problem, or condition under investigation | Effectiveness of prior infection with the SARS-CoV-2 Omicron BA.1 infection against reinfection with BA.2 and effectiveness of prior infection with the SARS-CoV-2 Omicron BA.2 infection against reinfection with BA.1. |
| Special considerations related to<br>Sex and gender | The effectiveness estimates were derived by comparing 1) incidence of BA.2 infection in the BA.1-infected cohort and uninfected-control cohort, and 2) incidence of BA.1 infection in the BA.2-infected cohort and uninfected-control cohort. Cohorts were exact-matched by sex to control for potential differences in the risk of exposure to SARS-CoV-2 infection by sex. |
| Age | Cohorts were exact-matched by 10-year age group to control for potential differences in the risk of exposure to SARS-CoV-2 infection by age. Nonetheless, with the young population of Qatar, our findings may not be generalizable to other countries where elderly citizens constitute a larger proportion of the total population. |
| Race or ethnicity group | Cohorts were exact-matched by nationality to control for potential differences in the risk of exposure to SARS-CoV-2 infection by nationality. Nationality is associated with race and ethnicity in the population of Qatar. |
| Geography | Individual-level data on geography were not available, but Qatar is essentially a city state and infection incidence was broadly distributed across the country's neighborhoods/areas. Cohorts were exact-matched by nationality to control for potential differences in the risk of exposure to SARS-CoV-2 infection by nationality. Qatar has unusually diverse demographics in that 89% of the population are international expatriate residents coming from over 150 countries from all world regions. |
| Other considerations | Individual-level data on co-morbid conditions were not available, but only a small proportion of the study population may have had serious co-morbid conditions. Only 9% of the population of Qatar are $\geq 50$ years of age (older age as proxy for co-morbidities). The national list of persons prioritized to receive the vaccine during the first phase of vaccine roll-out included only 19,800 individuals of all age groups with serious co-morbid conditions. Individual-level data on occupation were not available but matching by nationality may have (partially) controlled the differences in occupational risk, given the association between nationality and occupation in Qatar. |
| Overall representativeness of this study | The study was based on the total population of Qatar and thus the study population is broadly representative of the diverse, by national background, but young and predominantly male, total population of Qatar. While there could be differences in the risk of exposure to SARS-CoV-2 infection by sex, age, and nationality, cohorts were exact-matched by these factors to control for their potential impact on our estimates for effectiveness of prior infection with a SARS-CoV-2 Omicron sub-lineage (BA.1 or BA.2) against reinfection with the other sub-lineage. Given that only 9% of the population of Qatar are $\geq 50$ years of age and the limited proportion of the population with significant co-morbidities, our estimates of effectiveness may not be generalizable to other countries where elderly citizens constitute a larger proportion of the total population or where co-morbid conditions are prevalent. |

**Figure S1.**
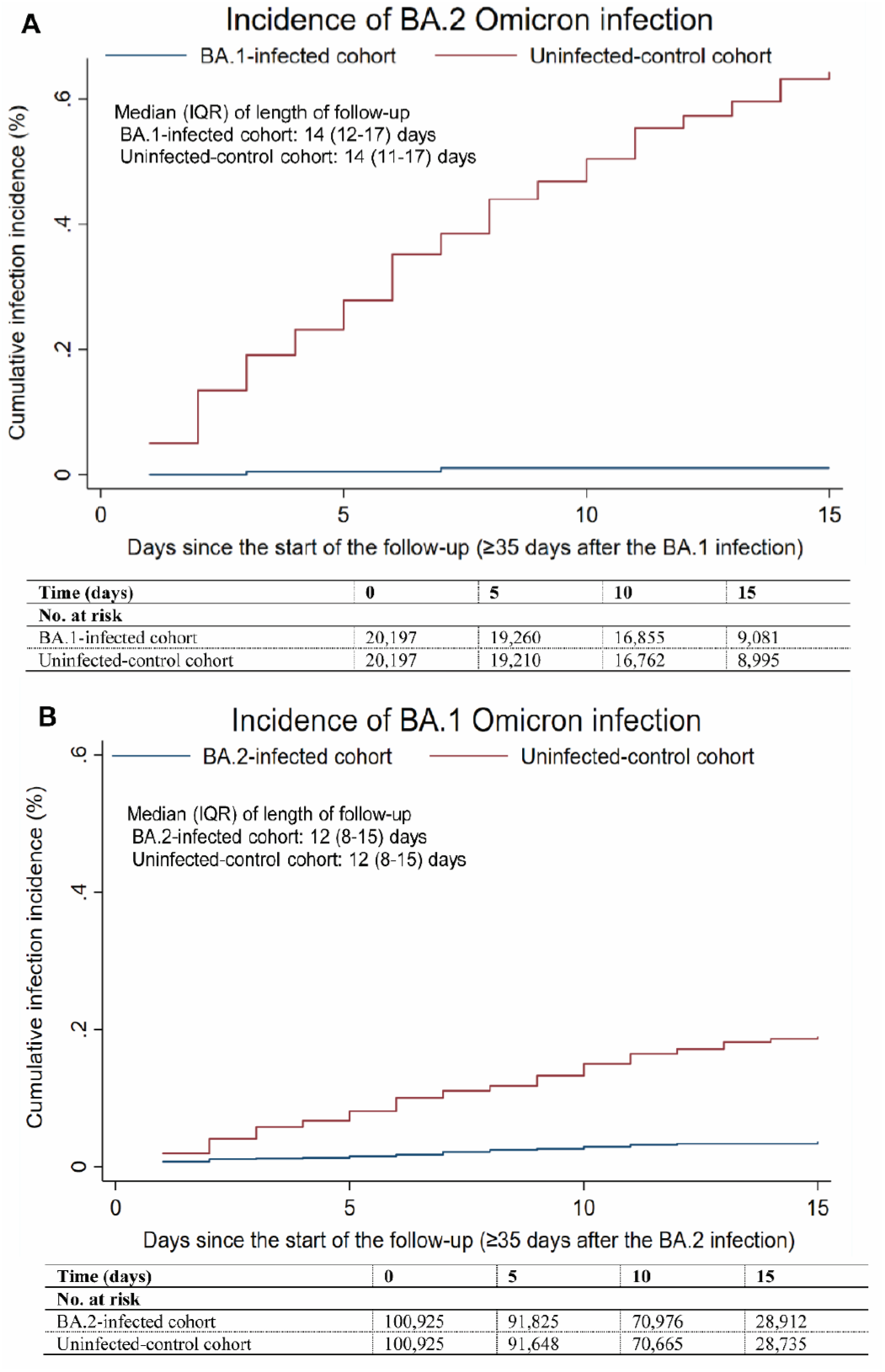
Sensitivity analysis. Cumulative incidence of A) BA.2 and B) BA.1 Omicron infections in the BA.1-against-BA.2 and BA.2-against-BA.1 studies, respectively.

